# Analysis of the COVID-19 epidemic in french overseas department Mayotte based on a modified deterministic and stochastic SEIR model

**DOI:** 10.1101/2020.04.15.20062752

**Authors:** Solym Mawaki Manou-Abi, Julien Balicchi

## Abstract

In order to anticipate a future trends in the development of the novel coronavirus COVID-19 epidemic started early at march 13, in the french overseas department Mayotte, we consider in this paper a modified deterministic and stochastic epidemic model. The model divides the total population into several possible states or compartment: susceptible (S), exposed (E) infected and being under an incubation period, infected (I) being infectious, simple or mild removed *R*_*M*_, severe removed (including hospitalized) *R*_*S*_ and death cases (D). The adding of the two new compartment *R*_*M*_ and *R*_*S*_ are driven by data which together replace the original *R* compartment in the classical SEIR model.

We first fit the constant transmission rate parameter to the epidemic data in Mayotte during an early exponential growth phase using an algorithm with a package of the software *R* and based on a Maximum Likewood estimator. This allows us to predict the epidemic without any control in order to understand how the control measure and public policies designed are having the desired impact of controlling the epidemic. To do this, we introduce a temporally varying decreasing transmission rate parameter with a control or quarantine parameter *q*. Then we pointed out some values of *q* to maintain control which is critical in Mayotte given the fragility of its health infrastructure and the significant fraction of the population without access to water.

## 1. Introduction

A novel coronavirus COVID-19 spread from the capital of the Hubei province in China to the rest of the country, then to most of the world and particularly the french overseas department Mayotte. Following the emergence of this novel coronavirus and its spread outside of China, Europe is now experiencing large epidemics. In response, France and many European countries have implemented unprecedented non-pharmaceutical interventions including case isolation of symptomatic individuals and their contacts, the closure of schools and universities, banning of mass gatherings and some public events, and most recently, widescale social distancing including local and national lockdowns of populations with all but essential internal travel banned. Also in response to the rising numbers of cases and deaths, and to maintain the capacity of health systems to treat as many severe cases as possible, France like European countries and other continents, have implemented some process measures to control the epidemic including in his overseas department outside the metropolitan France. The epidemic began slightly later in France, from February. To prevent further spread of COVID-19, all travel into Mayotte for tourism and family visits was prohibited from Friday, March 20. Only essential travel for selected individuals, such as residents returning home, professionals from essential services, and patients with health conditions was allowed to entry. Local authorities have advised all travelers arriving from the mainland France or areas affected by COVID-19 to self-isolate at home for 14 days and avoid contact with others. In addition, the government has urged the public to observe good personal hygiene, such as regular hand washing. Individuals who develop respiratory symptoms, such as cough, fever, and difficulty breathing, within 14 days of arrival into Mayotte are advised not to visit the doctor or emergency room directly, but to contact the SAMU (French Emergency Medical Services) immediately by dialing 15. Further international infected travellers of the virus was expected until March 29; but from March 30 the government has decided to cancel all flights both departure and arrival in Mayotte.

Many mathematical models of the COVID-19 coronavirus epidemic in China, USA, Italy and France have been developed, and some of these are listed in th following papers [**?, ?, ?, ?, ?, ?, ?, ?, ?**]. Mathematical models can be defined as a method of emulating real life situations with mathematical equations to expect their future behavior. In epidemiology, mathematical models are relevant tools in analyzing the spread and control of infectious diseases.

Based on the development and epidemiological characteristics of COVID-19 infection, a modified SEIR model is appropriate to study the dynamic of this disease. The population is partitioned into susceptible (S), exposed (E) infected and being under an incubation period, infected (I) being infectious, simple or mild removed *R*_*M*_, severe removed (including hospitalized) *R*_*S*_ and death cases (D).

It is well known that one of the most useful parameters concerning infectious diseases is called basic reproduction number. It can be specific to each strain of an epidemic model. Estimating the basic reproduction numbers for COVID-19 presents challenges due to the high proportion of infectious not detected by health systems and regular changes in testing policies, resulting in different proportions of infections being detected over time and between countries. Most countries so far only have the capacity to test a small proportion of suspected cases and tests are reserved for severely ill patients or for high-risk groups (e.g. contacts of cases appeared in Mulhouse,France). Looking at case data, therefore, gives a systematically biased view of trends. However, in France and other contries in Europe the basic reproduction numbers for COVID-19 is betwen 2 and 3 at the begining of the epidemic with an average around 2.5.

In our case, firstly we fit the basic reproduction number for COVID-19 to data in Mayotte at the begining of the epidemic in Mayotte. After that, we focused on the effects of the France government imposed public policies designed to contain this epidemic in his overseas department Mayotte. We found that the effective date compared to the prediction of the data without any control measure appears few days after.

Thus we assume that from this effective date where the control measure started, the transmission rate parameter *β* is time-depending and exponentially or log-exponentially decreasing with a control *c* or quarantine parameter *q* assume to be independent of the recovery rate since the COVID-19 is not actually curable.

Our model can thus be used to project the time-line of the model forward in time. We propose various scenarios on this control to show how the impact of strict respect of the government measure can lead to have a control on the capacity of the main hospital in Mayotte to manage severe cases according to the maximal number of staff and places available.

The government takes measures to track and quarantine people who have close contact with conrmed cases; it is quite obvious in theory but difficult in practice. In this paper we avoid to put in the model a compartment which account individuals through quarantine of infected individuals. We only show how controling the paramter *q* in the decreasing transmission because of the quarantine measure and public policies can lead to stop the epidemic.

The novelty of our paper compared to others mathematicals methods and model is based on our new model (**??**) applied to real data collected in Mayotte with initial conditions such that the initial exposed population with an infected individual which relies on the population density. We also consider different kind of parametrization of he transmission parameter assume to be a slowly varying decreasing piecewise deterministic function. This paper is organized as follows. Section 2 deals with some preliminaries and basis aspects of the deterministic SEIR model intended to clarify the computations and simulations. In section 3, we give a short description of Mayotte and the epidemic data. We present the numerical simulations of the epidemic including real data and predicted data in section 4. Section 5 deals with the stochastic modified model with various simulations and discussions about the relevance of stochastic model in small populations. Finally, we present in the appendix the deterministic method for computing *R*_0_ and a probabilistic extension of our seir model based the some early works of Etienne Pardoux and Tom Britton in [**?**].

## 2. Preliminaries and basis aspects of the deterministic SEIR model

A deterministic susceptible-exposed-infectious-recovered (SEIR) model for infectious diseases is developed here with the aim to make experimental simulations in next sections. Because the classical model assumes that the infected person’s incubation period is not infectious, this assumption is quite different from the infection characteristics of the new coronavirus infection. Therefore we will use a revised SEIR model to analyse and predict the trend of the outbreak. The model meets the following assumption.

(H1) The mortality rate *µ* induced by the disease is considered. This rate has been normalized compared to metropolitan France and according to the local reality in Mayotte.
(H2) The transmission method is person-to-person and after a short infectious period individual becomes permanently immune.
(H3) Once exposed, individuals go through a latent period 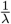 of approximately 6 days after which they become infectious for a period 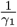 that is estimated to be 8 days for slight or mild infectious cases and for a period 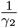 for severe infectious cases that is estimated to be 21 days.
(H4) We assume that there exists a proportion *p* for slight or mild infections.

The model divides the total population into six categories:

- Susceptibles **S** not yet infected but a risk of being infected
- Exposed **E** infected but not yet be infectious, in a latent period; Infectious
- Confirmed **I** to be infectious (with infectious capacity and not yet be quarantined) and contain reported and unreported cases.
- Removed with mild infections **R**_*M*_, Removed **R**_*S*_ with severe infections and **Death** death cases.

Define *S*(*t*) *E*(*t*), *I*(*t*), *R*_*M*_ (*t*), *R*_*S*_(*t*) and *D*(*t*) as the number of susceptible, exposed, infectious, removed with mild infections, severe infections including hospitalization and death cases in the population at time *t*, respectively.

Consider a time interval [*t, t* + *dt*] where the small change *dt* represents the length between the two points at which measurements are taken. Let *β* denote the disease transmission coefficient. This can be though as the rate at which each infectious individual makes potentially infections contacts with each other individual, where a potential infectious contacts will transmit the disease if the contact is made by an infectious individual with a susceptible individual. It is the product of the contact rate and the probability of infection given a contact per unit time.

The number of new infections in the time interval [*t, t* + *dt*] is

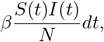

where *N* is the total population size at the beginning date *t*_0_ of the epidemic. Given initial conditions *S*(0) = *N*, initial average of exposed individuals *E*_0_ *>* 0 and *I*(0) = *I*_0_ *>* 0, the model consists of the following system of ordinary differential equations:

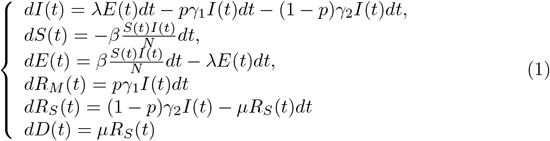

The total population size is denoted by *N*(*t*) = *S*(*t*) + *E*(*t*) + *I*(*t*) + *R*_*M*_(*t*) + *R*_*M*_(*t*) + *D*(*t*) and satisfies 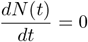, so that the population size remains constant.

### Remark 2.1

*It is worth to mentionned that the initial average of exposed individuals in front of an infectious individuals E*_0_ *is not E*(0).

**Figure 1.**
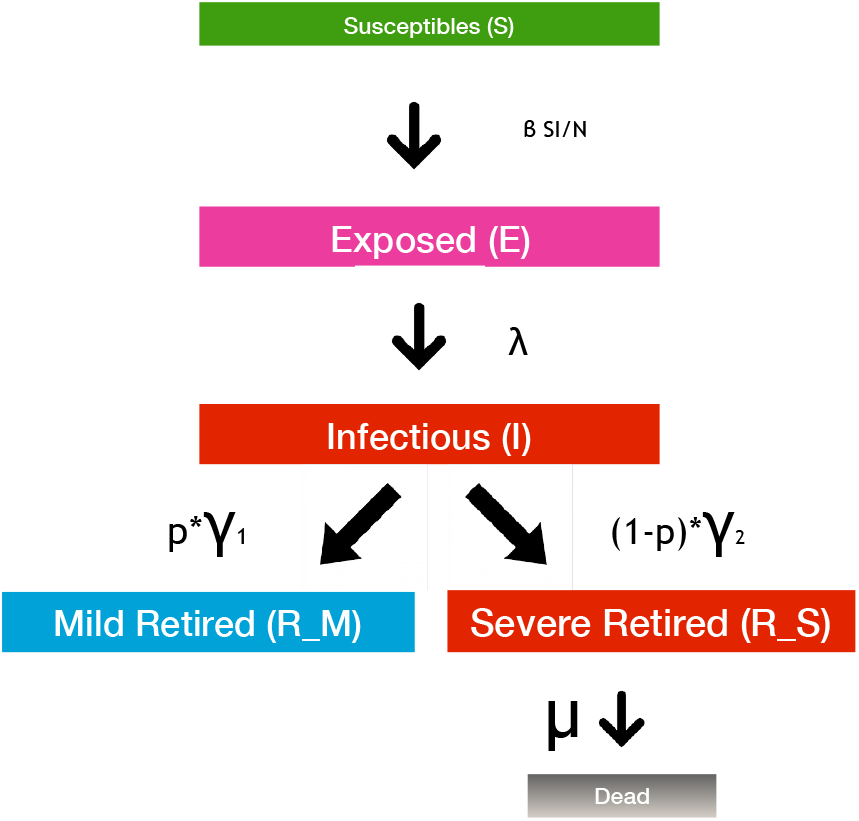
Compartments of the SEIR model.

One can also look, by this model, to keep track of the cumulative number of Covid-19 cases from the time of onset of symptoms for *C*(*t*) which is not a compartent and defined as follows:

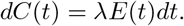

With contact tracing, a proportion of individuals exposed to the virus is quarantined.

### Remark 2.2

*One can assume that some international infectious travelers individuals y*(*t*) *negligible in front of the total population size S*(*t*) (*S*(*t*) + *y*(*t*) *∼S*(*t*)) *but not negligible in front of Infectious individuals I*(*t*) *subsequently travel at time t and are eventually detected in their destination to Mayotte*.

**Figure 2.**
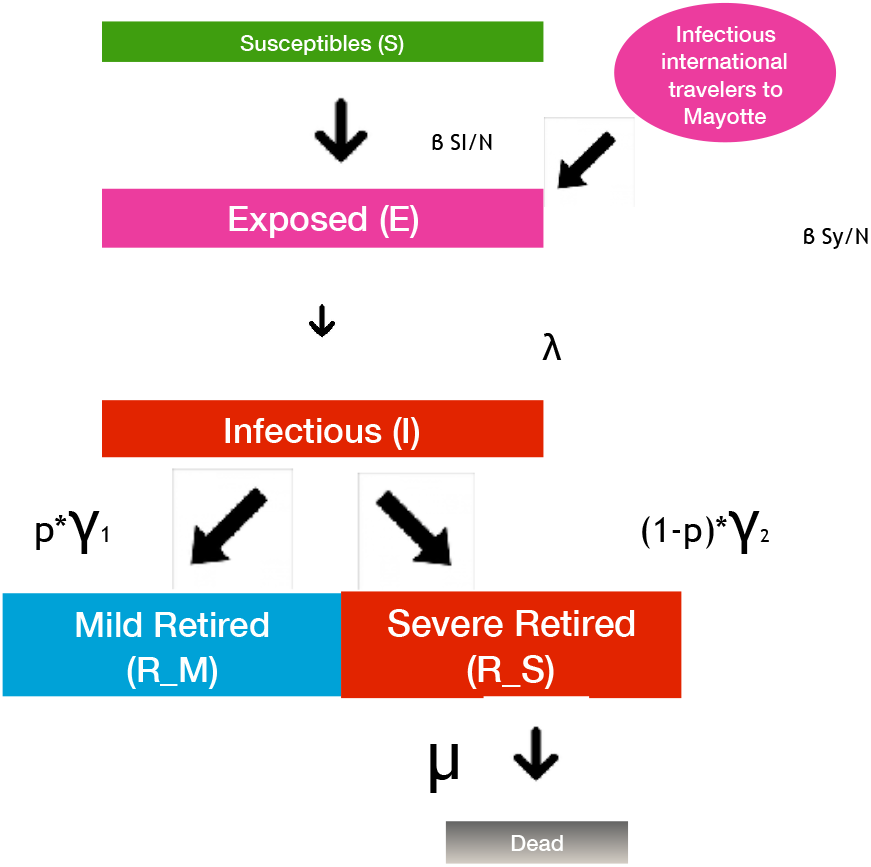
Compartments of the SEIR model including infectious travellers.

Hence this leads to the new equations of the model:

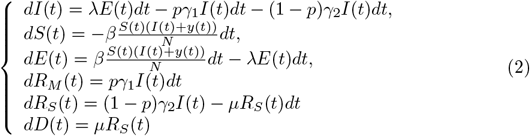

The dynamic of new exposed travellers *y*(*t*) can be thought as follows:

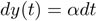

where *α* is the average number of imported exposed individuals.

The following table lists the parameters of the model we’ll use for numerical simulation and estimates. These estimates are often wide due to wide variations from one infection to another. Only the parameter *β* will be estimated.

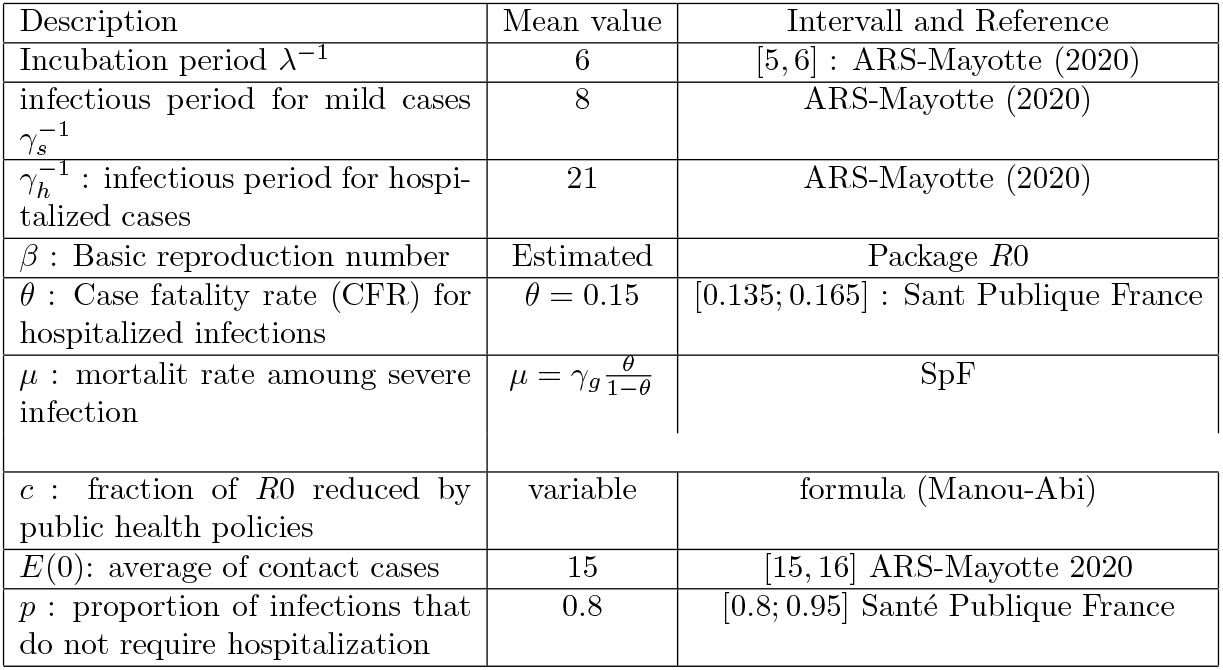

| Description | Mean value | Intervall and Reference |
| --- | --- | --- |
| Incubation period $\lambda^{-1}$ | 6 | [5, 6] : ARS-Mayotte (2020) |
| infectious period for mild cases $\gamma_s^{-1}$ | 8 | ARS-Mayotte (2020) |
| $\gamma_h^{-1}$ : infectious period for hospitalized cases | 21 | ARS-Mayotte (2020) |
| $\beta$ : Basic reproduction number | Estimated | Package $R_0$ |
| $\theta$ : Case fatality rate (CFR) for hospitalized infections | $\theta = 0.15$ | [0.135; 0.165] : Sant Publique France |
| $\mu$ : mortalit rate amoung severe infection | $\mu = \gamma_g \frac{\theta}{1-\theta}$ | SpF |
| $c$ : fraction of $R_0$ reduced by public health policies | variable | formula (Manou-Abi) |
| $E(0)$: average of contact cases | 15 | [15, 16] ARS-Mayotte 2020 |
| $p$ : proportion of infections that do not require hospitalization | 0.8 | [0.8; 0.95] Santé Publique France |

### Remark 2.3

*In the above model, we only consider constant-valued parameters. However, many of these quantities actually vary with time. For instance in the absence of vaccination and lack of effective treatment, the only way to influence the disease development is to act on the basic reproduction number R*_0_ *through for instance β; that is to decrease the value of this parameter. One can then thing naturally about a decreasing time varying transmission coefficient β*(*t*) *that allows the effect of control interventions so that to explore how the effective reproduction number R*_0_(*t*) *decrease in time*.

According to this remark, the previous system of ordinary differential equations becomes:

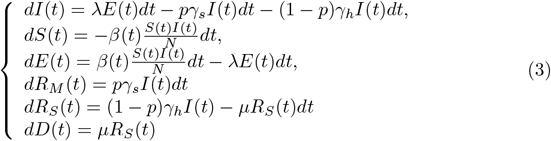

This is a realistic model that can be used by epidemiologists to study the dynamics of the diseases and the effects of control interventions. The basic reproduction number *R*_0_ is defined as the average number of secondary cases generated by a primary case over his/her infectious period when introduced into a large population of susceptibles individuals [**?**]. Note that the constant *R*_0_ thus measures the initial growth rate of the epidemic and for the model above it can be shown (see the next section) that

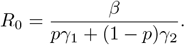

Furthermore Chowell et al. [**?**] define the time-dependent effective reproductive number

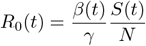

of secondary cases per infections cases at time *t*. One can see that if *S*(*t*) *∼ N* it follows that 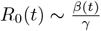 is a function proportional to the time varying transmission coefficient *β*(*t*). The time point at which *R*_0_(*t*) assumes values smaller that 1 indicates when control measures have becomes effective in controlling the epidemic. The intervention strategies to control the spread of Covid include surveillance, placement of suspected cases in quarantine for two weeks (the maximum estimated length of the incubation period), education of hospital personnel and community members on the use of strict barrier nursing techniques (i.e. protective clothing and equipment, patient management), and the rapid burial or cremation of patients who die from the disease. Reducing exposure is one of the effective measures to control the spread of disease. The public raises awareness of precaution and takes fewer trips or wears a mask in the presence of a disease outbreak.

In order to account for the control intervention we assume, in our model, that the transmission rate function *β*(*t*) is a decreasing piecewise linear continuous function up to the time point *t*_0_ when the control measures are introduced. Assumes that a fraction *c∈* [0, 1) of new infections is reduced during a period *T* = *t* − *t*_0_. Then we can write:

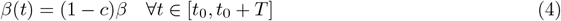

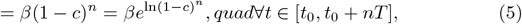

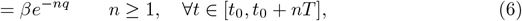

where *q* =− ln(1− *c*)≥0 is a kind of quarantine control parameter. Thus, we assume in this paper that *β*(*t*) takes the following form:

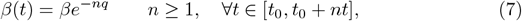

where *n* equal to the period where the control parameter relied for example one can assume that the percentage of a control measure is maintened about a 7-day period. This will be the approach we use for simulations in the paper.

Note that such kind of parametrization of *β*(*t*) was already consider in [**?**] and [**?**]. Let us point out that the geometry of the likelihood function can not permit identification of this quarantine parameter *q* as its estimator is correlated to *β*.

Our way for the transmission rate is a more parsimonious parametrization and it does not affect *γ* the recovery rate unless the disease is curable, which is not the case for Covid-19 actually. Now, let us discussed about the importance of *c* and *q*.

- If *c* = 0 (*q* = 0) one can see that *β*(*t*) = *β* which means that 0% of new infections are reduced so that there is no effort to contain new infections which mean that the control intervention failed.
- Note that in the time interval [0, *t*−*t*_0_] if we consider *q* = 0.2 (*c∼*0.18) and *β* = 0.422 (see the next section for more details ont his value) this is equivalent to a higher level of control measures since this mean that after time *T* the control measure started, approximately 80% of new interventions are reduced. One can then thing on 1−*p* as the percentage of succefull intervention measures during the major public policies process introduced to avoid new infections.
- If a fraction *c* of new infections is reduced during every period *T* = *t*−*t*_*\**_ then the epidemic will go to extinction after *nT* time where *n* is the interger part of

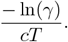

In the spirit of the above parametrization, we will consider according to observation in real data where we think that there is a slight increasing in a sequence of change times *t*_1_, …*t*_*K*_ and followed by a slowly varying exponential decrease, a piecewise linear continuous transmission coefficient *β*(*t*) in the form:

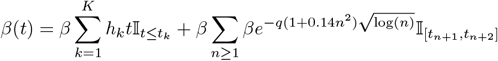

where *β* = *β*(0) at the begining of the epidemic and *n* = 1, 2 denoting the change of one week to another.

## 3. About the french overseas department M ayotte and epidemic Data

Mayotte, is an island and overseas department of France located in the northern Mozambique Channel in the Indian Ocean and the coast of Southeast Africa, between northwestern Madagascar and northeastern Mozambique. It is also located between the two southeasternmost islands of the Comoros archipelago. Mayotte is made up of one main island (also called Grande Terre), one smaller island called Petite Terre which lies about 1.5 miles east of the main island and islets. The capital, Mamoudzou, is located on the eastern coast of the main island and in the neighbouring smaller island of Petite-Terre, the Dzaoudzi International Airport is located. The territory is also known as Maore, the native name of its main island, especially by advocates of its inclusion in the Union of the Comoros. Mayotte has a total land area of about 144 square miles or 374 square kilometres (sq. km), and with its almost 285000 people according to official estimates, is very densely populated at 747 inhabitants per sq.km. Mayotte is the poorest department of France. However, many immigrants from other Mozambique Channel nations enter Mayotte illegally, as it is one of the most prosperous regions in the area.

From 1976 into the 21st century, Mayotte had a special status with France as a territorial collectivity, conceived as being midway between an overseas territory and an overseas department. Its status was changed to a departmental collectivity in 2001 and then to overseas department in 2011. Mayotte is represented in the French National Assembly by a deputy and in the French Senate by two senators. It is administered by a French-appointed prefect and an elected Departmental Council. The judiciary is modeled on the French system. Although, as a department, Mayotte is now an integral part of France, and despite being in France, the vast majority of Mayottes population do not speak French as a first language, but a majority of the people 14 years and older report in the census that they can speak French (with varying levels of fluency). The majority language is Shimaore, a language related closely to those found in the neighbouring Comoros islands. Kibushi, a Malagasy language, is the second most widely spoken. Both have been influenced by Shimaore and the vast majority of the population is Muslim.

As concerned the land, note that a volcanic mountain range forms a north-south chain on Mayotte island, with summits from about 1, 600 to 2, 000 feet (500 to 600 metres) in elevation. Protected waters for shipping and fishing are created by surrounding coral reefs some distance from the shore. Mayotte age structure is as follows: 46.6% of the population is under age 15, 51% of the population between 15 and 64 years old and 2.4% of the population is older than 65. As we can see the Mayotte population is young. This type of age structure is common for developing countries with high birth and death rates. Relatively short life expectancy, as well as low level of education and poor health care.

**Figure 3.**
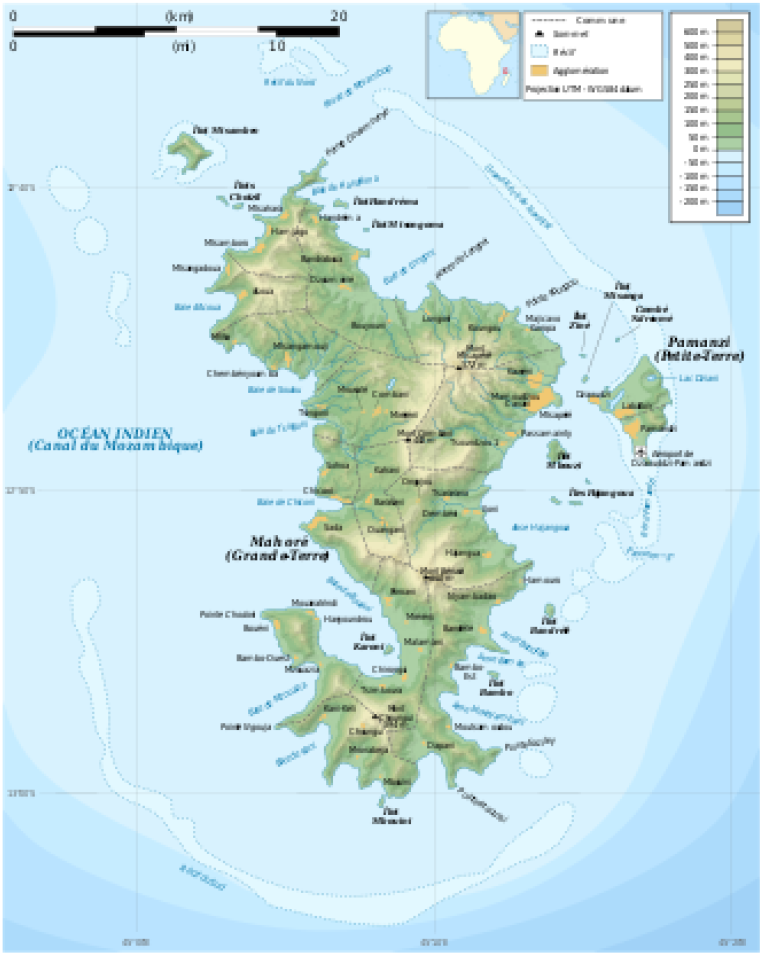
A map of Mayotte.

### 3.1 Epidemic Data process in Mayotte

Mayottes Regional Health Agency and local authorities confirms the first case of new coronavirus infection on march 14 but reported on march 13, an imported case. He was an individual who traveled from the metropolitan France and back to Mayotte three days ago. Subsequently, two others imported case was reported few days later but arrived at the same moment like the first one. According to this information we choose the initial number of infected people *I*(0) = 3.

But 5 others imported case was confirmed on march 20. After that 8 others imported on March 26. Again three others imported cases were confirmed from march 27. Thus a total of 17 imported cases were confirmed two week days later.

One can thus think that almost 1 imported case is introduced per day after march 13 until march 28. But since March 29, the authorities of Mayotte department and France have decided to close and cancel all flights to and from Mayotte, which therefore limits the introduction of new imported cases but cannot prevent the spread of the virus.

In this case and according to Remark **??**, we can assume that among susceptibles individuals a negligible fraction of international infectious individuals subsequently travel and are detected in their destination to Mayotte. Thus, according to the model (**??**) one may take *y*(*t*) = *t* as a variable number *y*(*t*) of infectious international travellers to Mayotte at time *t*. Clearly, this is reasonable according to the epidemic history data in Mayotte. Hence for *t∈* [*t*_0_, *t*_*c*_], this leads to the following new equations of the model:

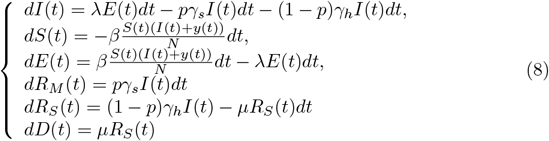

where *t*_0_ = *march*13 and *t*_*c*_ = *march*29. But for *t* ≥ *t*_*c*_ +1 we assume that *y*(*t*) = 0.

We obtain information and data from the regional french agency of health called ARS of Mayotte where the second author of this paper works.

| Dates | imported cases |
| --- | --- |
| from march 13 | 3 |
| from march 14 to 27 | + 14 |
| from march 28 to nowadays | + 0. |

The regional french agency of health (ARS) of Mayotte according with the French Public Heath (SpF) started to release a daily bulletin about COVID-19 infections in Mayotte. According to ARS Mayotte and SpF, the probable contact number with a confirmed case is around 15−16 so we take for our model *E*_0_ = 15 at the begining of the epidemic.

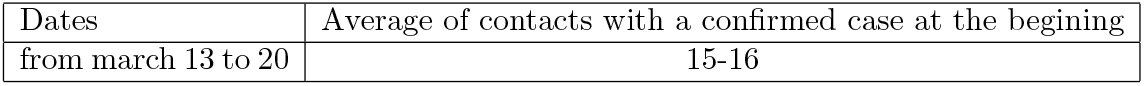

| Dates | Average of contacts with a confirmed case at the beginning |
| --- | --- |
| from march 13 to 20 | 15-16 |

More informations can be found at the following website of the Mayotte local authorities: http://www.mayotte.gouv.fr and www.mayotte.ars.sante.fr in Coronavirus COVID-19: Point de situation. The following table presents the data observed in Mayotte.

Note that data information includes the cumulative number of reported (confirmed), imported, hospitalized cases including critical cases. For more informations about the collection of the data, we refer the reader to the following website of the Mayotte local authorities: http://www.mayotte.gouv.fr or www.mayotte.ars.sante.fr in Coronavirus COVID-19 Point de situation.

It should be noted that in fact it is the data from the previous day which is presented the next day.

**Table 1.** Covid cases in Mayotte from march 13 to 19

| Date | march 13-14 | march 15 | march 16 | march 17 | march 18 | march 19 | march 20 |
| --- | --- | --- | --- | --- | --- | --- | --- |
| Confirmed cases | 1 | 1 | 1 | 2 | 3 | 4 | 6 |
| incidence cases | 1 | 0 | 0 | 1 | 1 | 1 | 2 |
| imported cases | 1 | 1 | 2 | 2 | 3 | 3 | 3 |
| hospitalized cases | 0 | 0 | 0 | 0 | 0 | 0 | 1 |
| critical cases | 0 | 0 | 0 | 0 | 0 | 0 | 1 |
| cured cases | 0 | 0 | 0 | 0 | 0 | 0 | 0 |
| death cases | 0 | 0 | 0 | 0 | 0 | 0 | 0 |

**Table 2.**
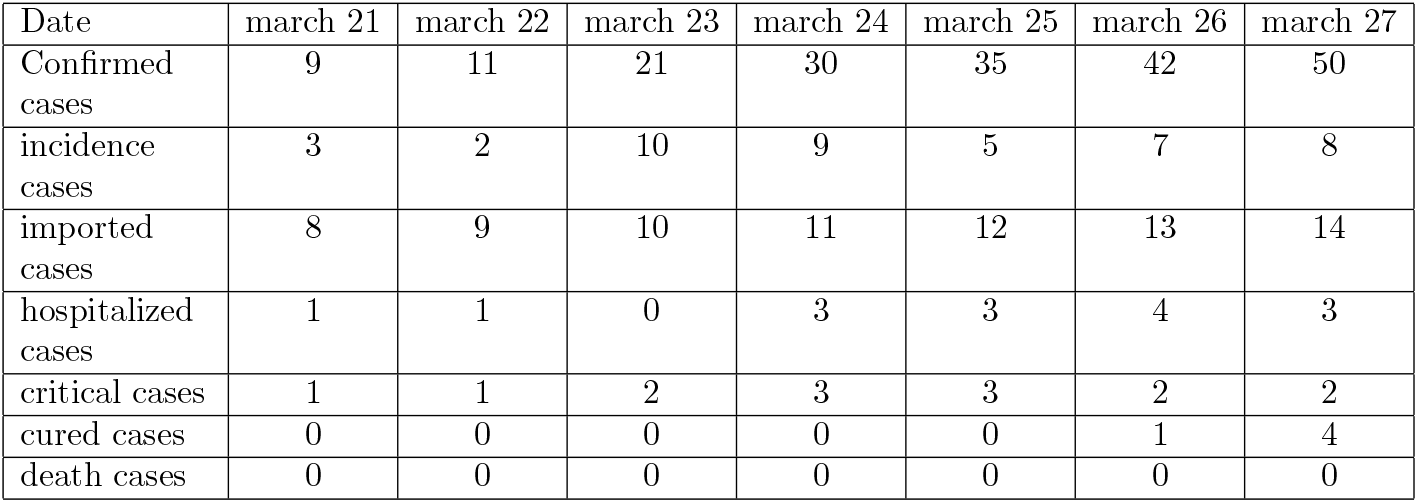
Covid cases in Mayotte from march 20 to 26

**Table 3.** Covid cases in Mayotte from march 27 to april 3

| Date | march 28 | march 29 | march 30 | march 31 | april 1 | april 2 | april 3 |
| --- | --- | --- | --- | --- | --- | --- | --- |
| Confirmed cases | 63 | 74 | 82 | 94 | 101 | 116 | 128 |
| incidence cases | 13 | 11 | 8 | 12 | 7 | 15 | 12 |
| imported cases | 17 | 17 | 17 | 17 | 17 | 17 | 17 |
| hospitalized cases | 3 | 8 | 10 | 10 | 11 | 13 | 14 |
| critical cases | 2 | 2 | 3 | 2 | 2 | 3 | 3 |
| cured cases | 4 | 8 | 10 | 10 | 10 | 10 | 13 |
| death cases | 0 | 0 | 0 | 1 | 1 | 2 | 2 |

**Table 4.**
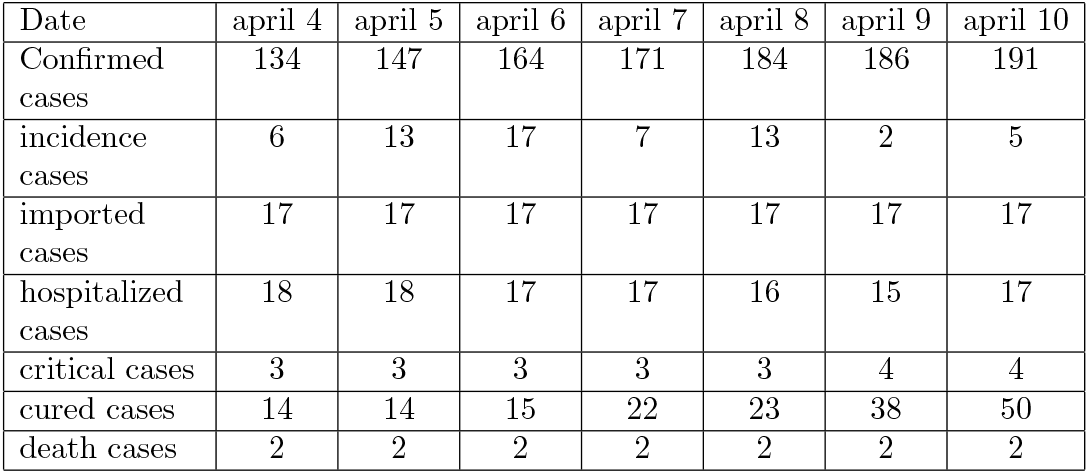
Covid cases in Mayotte from april 4 to 10

| Date | april 4 | april 5 | april 6 | april 7 | april 8 | april 9 | april 10 |
| --- | --- | --- | --- | --- | --- | --- | --- |
| Confirmed cases | 134 | 147 | 164 | 171 | 184 | 186 | 191 |
| incidence cases | 6 | 13 | 17 | 7 | 13 | 2 | 5 |
| imported cases | 17 | 17 | 17 | 17 | 17 | 17 | 17 |
| hospitalized cases | 18 | 18 | 17 | 17 | 16 | 15 | 17 |
| critical cases | 3 | 3 | 3 | 3 | 3 | 4 | 4 |
| cured cases | 14 | 14 | 15 | 22 | 23 | 38 | 50 |
| death cases | 2 | 2 | 2 | 2 | 2 | 2 | 2 |

**Table 5.** Covid cases in Mayotte from april 11 to 17

| Date | april 11 | april 12 | april 13 | april 14 | april 15 | april 16 | april 17 |
| --- | --- | --- | --- | --- | --- | --- | --- |
| Confirmed cases | 196 | 203 | 207 | 217 | 221 | 233 | 245 |
| incidence cases | 5 | 7 | 4 | 10 | 4 | 12 | 12 |
| hospitalized cases | 20 | 20 | 22 | 20 | 15 | 18 | 22 |
| critical cases | 3 | 3 | 3 | 3 | 3 | 4 | 6 |
| cured cases | 59 | 67 | 69 | 69 | 77 | 94 | 117 |
| death cases | 3 | 3 | 3 | 3 | 3 | 3 | 4 |

## 4. Illustration and Numerical simulations of the epidemic COVID-19 in Mayotte

The following picture illustrate real numbers of cumulative confirmed, cured, hospitalized and death cases from march 13 to april 17.

**Figure 4.**
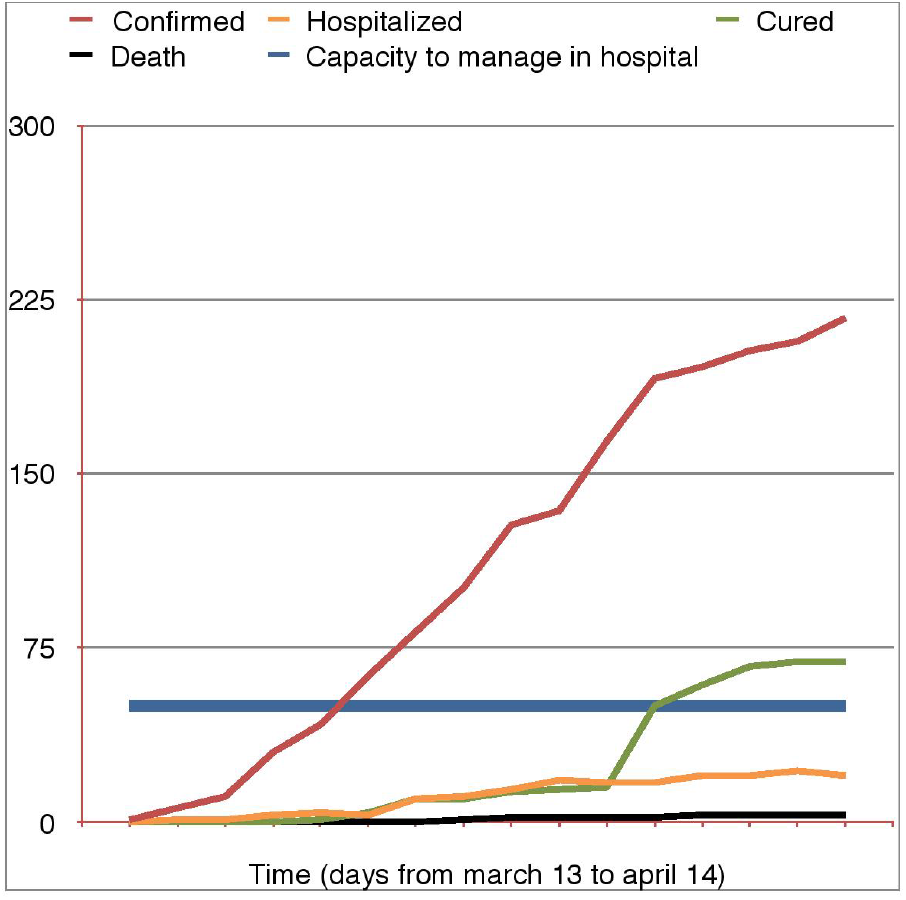
Covid19 in Mayotte: cumulative con_rmed, cured, hospitalized and death cases from march 13 to april 14.

### 4.1 Estimation of the basic reproduction number *R*_0_ in the begining of the epidemic

The Reproduction number *R*_0_ may be estimated at different times during an epidemic. Some importants review found that the estimated mean *R*_0_ for COVID-19 is in [2, 3] with a median of2.5 which is considerably higher than the SARS estimate or the influenza.

These estimates of *R*_0_ depend on the estimation method used as well as the validity of the underlying assumptions. Due to insufficient data and short onset time, current estimates of *R*_0_ for *COVID* − 19 are possibly biased. However, as more data are accumulated, estimation error can be expected to decrease.

Methods for estimating the basic reproduction number at the beginning of the outbreak and the time-dependent reproduction number of secondary cases at any time during the outbreak, are usually bases on the following methods, but all required some hypotheses in order to apply the methods.

For instance, the Maximum Likewood (ML) that we are interested in this paper as an estimator according to [**?**] and [**?**] propose extensions and options implemented in the software *R*. Others methods like Exponential growth rate (EG) and Sequential Bayesian method also exists.

Note that the ML methods require the user to select the time period over which growth is exponential and to determine the serial intervall meaning the duration between symptoms onset of the primary cases and symptoms onset of the secondary cases in a transmission chain.

In the case of *Covid* − 19 in Mayotte, the epidemic curve can be analyzed the first week from the case on march 15 to march 21 where over consecutive time units and a generation time interval with mean 2 days and standard deviation 0.5 day. To estimate the early dynamics of *Covid* − 19 transmission in Mayotte, we fitted the value of the serial interval with a gamma distribution with mean 2 and standard deviation 0.5.

By using the *R*_0_ Package, we estimate that the basic reproduction number for the epidemic in Mayotte

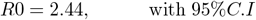

We presented the code using the *R*_0_ Package in the following lines.

**Table 6.**
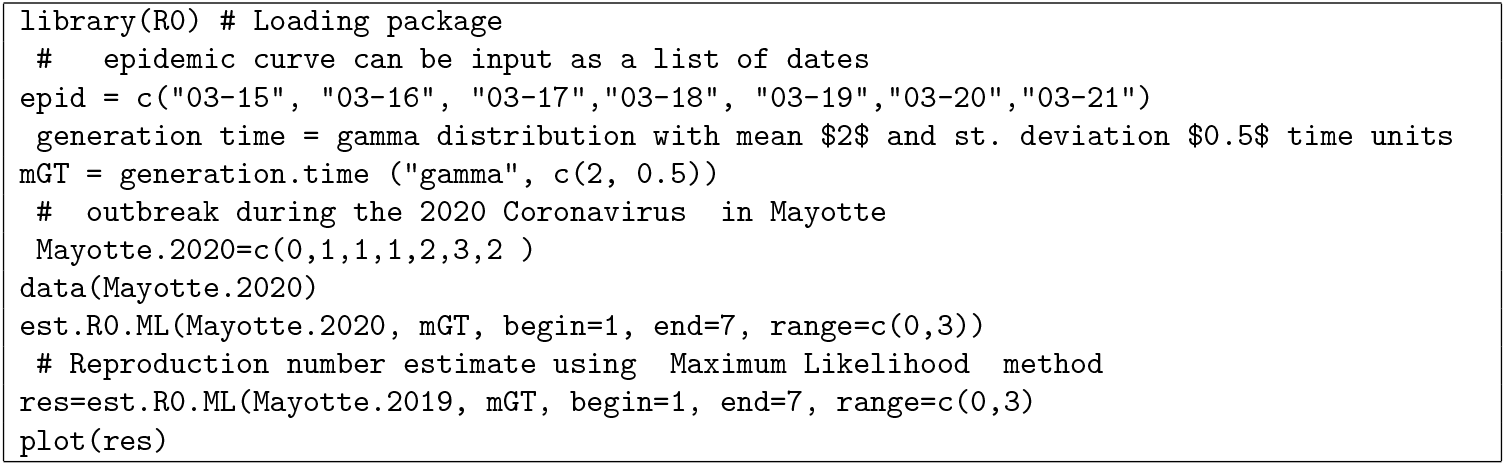
Typical session code for the basic reproduction number

### 4.2 Prediction of the Covid-19 in Mayotte

Now, we are able to apply the preceding compartmental model for the prediction of the *Covid*−19 in Mayotte. According to our assumptions on the infectious period we have

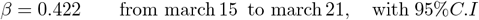

Assume that there was any intervention control, of course this is not true beacause from march 16, the french government imposed public policies designed to contain this epidemic in all departments including overseas department, for instance in Mayotte. The following table illustrative cumulative confirmed compared to predicted data with our mathematical model.

**Table 7.** Reported Covid and predicted cases in Mayotte from march 13 to april 10

| Dates | march 13 | march 20 | march 27 | april 3 | april 10 |
| --- | --- | --- | --- | --- | --- |
| Confirmed cases | 1 | 9 | 50 | 128 | 196 |
| Predicted without control measure | 1 | 13 | 43 | 109 | 292 |
| Predicted severe cases | 1.5 | 5 | 12 | 22 | 34 |
| Confirmed death cases | 0 | 0 | 0 | 2 | 2 |
| Predicted death cases | 0 | 0 | 0 | [[1,2]] | 3 |

#### Remark 4.1

*Note that during in this period the model predict that at most two hospitalized individual will died. This was the case at april 10 in Mayotte*.

### Discussion

Comparing model predictions with observed confirmed cases reported in Mayotte, we found that, from april 3 to 10 the model predicted almost two times higher cases than those were reported. Thus, we deduce that the control measures and public policies really started around April 1 instead of March 16 the official date.

The following picture illustrate the number of cumulative predictions cases in Mayotte as weel as the pic and the end of the epidemic without any control measure.

#### Discussion

Without any control measure, we can see that the pic will be attained around June 25 an the extinction of the epidemic can be esperate at the end of august.

##### Remark 4.2

*Note also that the cancelation of travel restrictions on the peak time and peak value has an effect on the peak time and peak value*.

## 5. Prediction when varying the quarantine control parameter *q*

Now let us assume that from march 20, the transmission rate parameter is time-depending, does not affect the recovery rate unless the disease is curable, which is not the case for Covid-19 actually and exponentially decreasing as described in **??** with a control quarantine paramter *q*. This should be actually the case since the control measure started at this time.

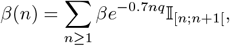

with *n* = 1 means march 20 and *n* = 2 means march 27 and so one.

**Figure 5.**
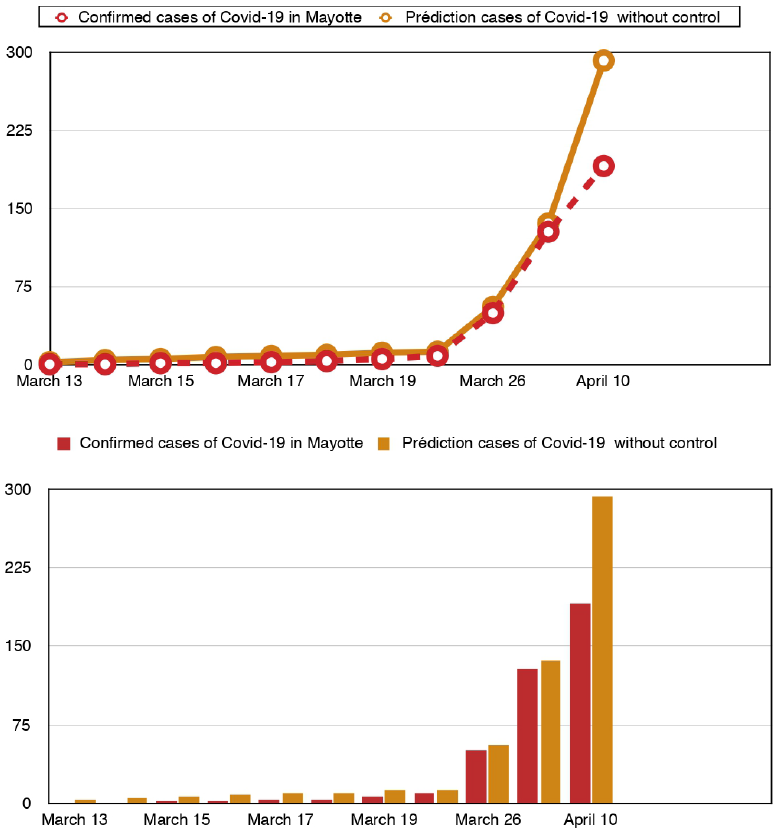
Predictions cases of Covid19 in Mayotte from march 26 to april 10 without any public policies.

The following table show the evolution of the epidemic when the control parameter *q* = 0.2 under the following time varying function

### Discussion

Under control measure with a control paramter *q* = 0.2, one observed that the pic is attained later around July 17 with a maximum of infectious people less that the one with no control measure.

#### Discussion

Now, let us remark that at this stage with the exponential decreasing parameter *β* if *q* = 0.5 every 7 days one can observe a rapide extinction of the epidemic but with *q* = 0.05 or *q* = 0.07 every 7 days the extinction come later. At this stage, we hink that a different parametrization need to be introduced. So one would like to imagine a slowly varying parameter in the form

**Figure 6.**
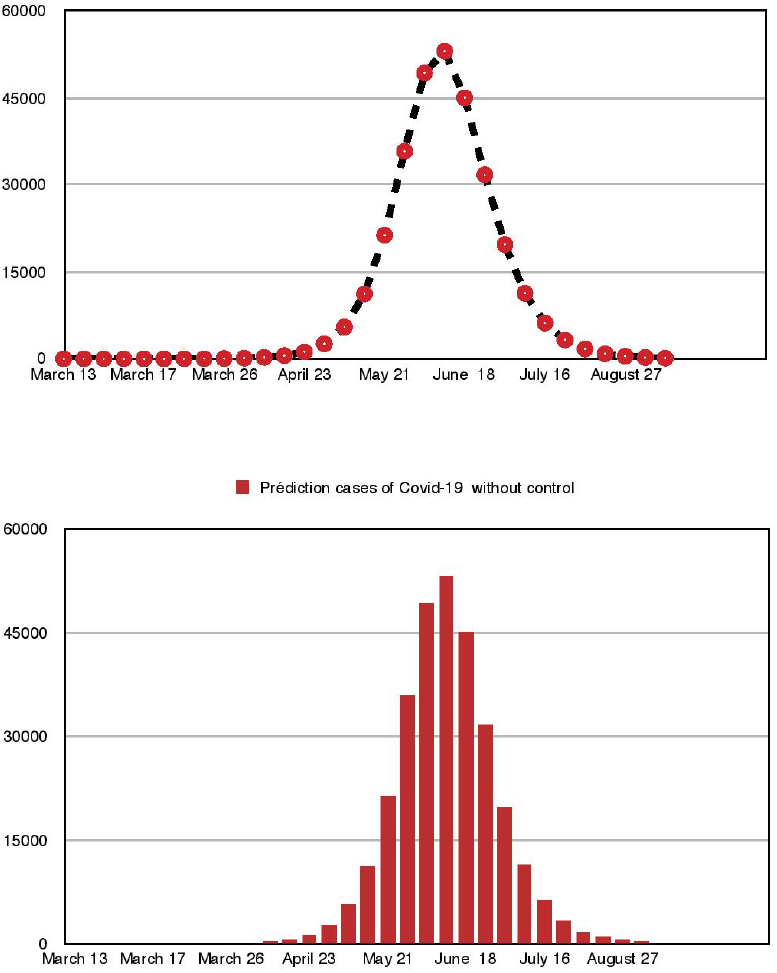
Predictions cases of Covid19 in Mayotte without any public control.

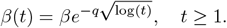

But since we already observed that the control measures and public policies really started around April 1 instead of March 16 the official date, we will assume that there was an increasing of the transmission parameter *β*(*t*) during the second week (from March 20 to 26 and from March 26 to April 3) and after that the decrease phase started

**Figure 7.**
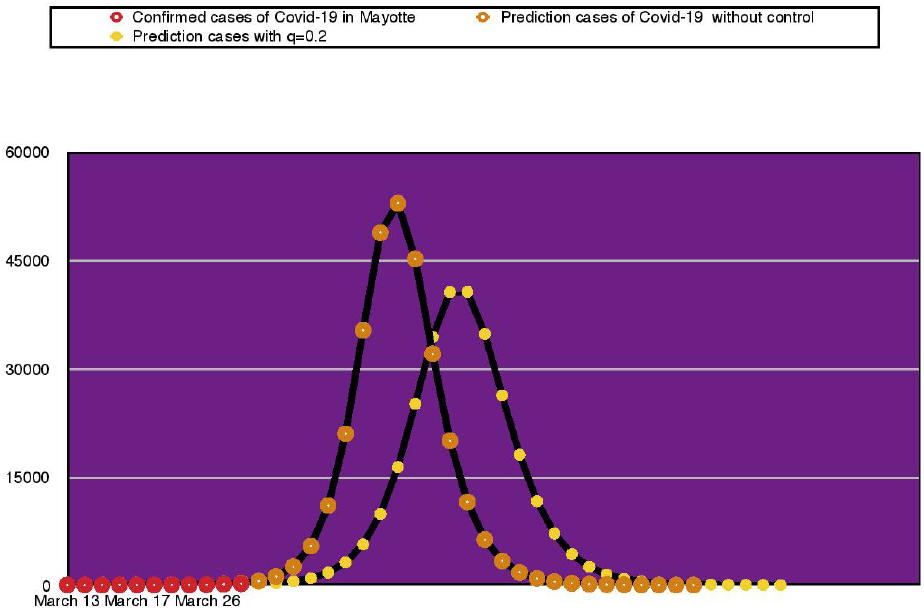
Predictions cases of Covid19 in Mayotte with a control paramter q = 0:2.

Now we assume in the rest of the prediction scenarios that there was a slowly varying in transmission on the form

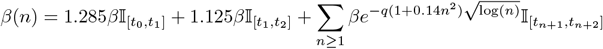

where *β* = *β*(0) at the begining and

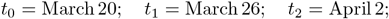

**Figure 8.**
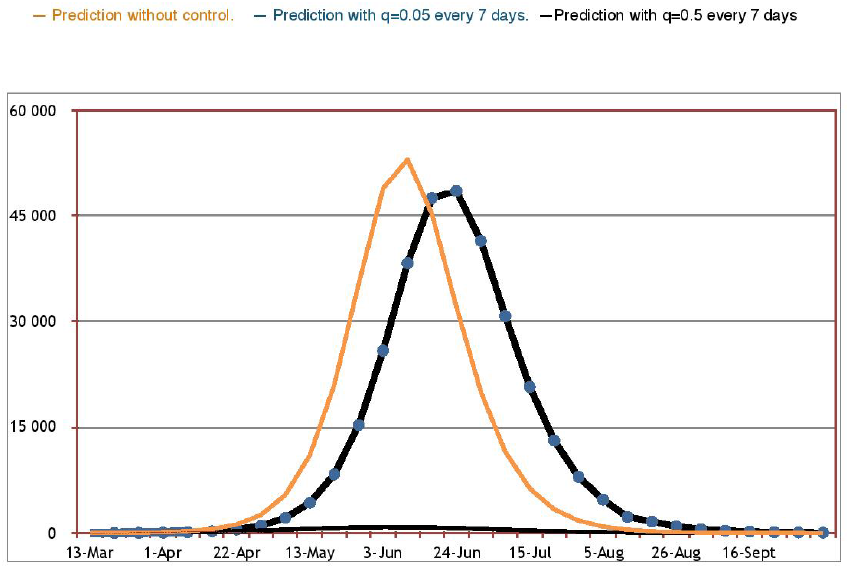
Predictions cases of Covid19 in Mayotte with various control parameter.

and *n* = 1, 2 denoting the change of one week to another.

Let us see what becomes the simulation with a specific choosen quantine parameter *q* = 0.1 compared with reported Covid-19 cases in Mayotte. To examine the possible impact of enhanced interventions on disease infections, we plotted the number of cumulative infected individuals *I*(*t*) and the predicted cumulative number of reported cases with varying the quarantine parameter *q*. This analysis shows that incresing the parameter persistently decreases the peak value but may either delay or bring forward the peak, as shown in the above pictures.

##### Remark 5.1

*Note that, enhancing quarantine and isolation following contact tracing and reducing the contact rate can signicantly lower the peak and reduce the cumulative number of predicted reported*

**Figure 9.**
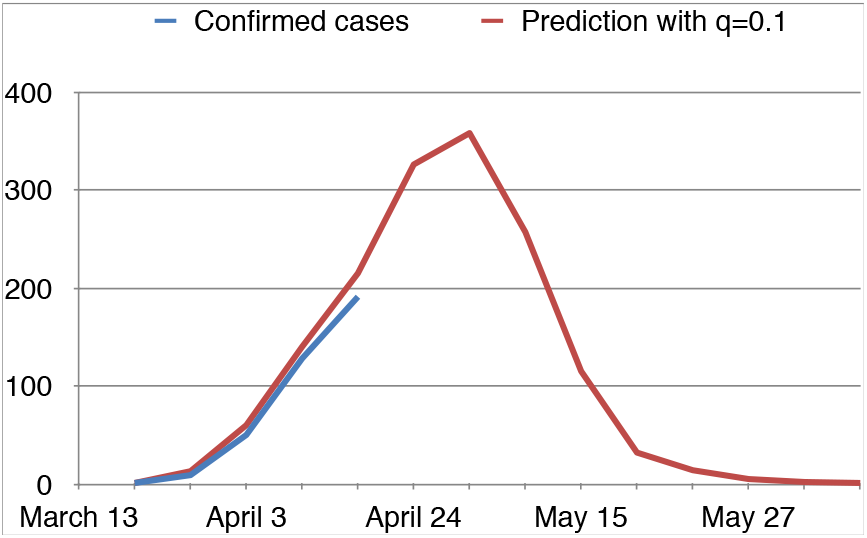
Predictions cases of Covid19 in Mayotte with the new control parameter.

### 5.1 Discussion about the relevance about stochastic models

At this stage, let us point out the fact that a stochastic pertubation is necessary since in general, susceptible individual does not have the same probability of being infected and depends on many factors depending on the behavior of individuals which changes over the course of the epidemic for instance the confinement will changes the habit. Therefore suppose that some stochastic environmental factor acts simultaneously on each individual in the population so that *β* changes to a a stochastic process (*β*(*t*))_*t*≥0_ such that

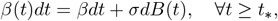

where *B* is a standard brownian motion. Hence the number of potential infectious contacts that a single infected individual makes with another individual during a period *dt* is normally distributed with mean *βdt* and variance *σ*^2^*dt*. Hence

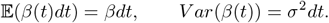

As *V ar*(*β*(*t*))*→*0 when *dt→*0, this is reasonable model for biologist and it is a well-established way of introducing stochastic environmental noise into biologically realistic population dynamic models. To motivate the above assumption on *β*, let us we argue as follows. Suppose that thenumbers of potentially infectious contacts between an infectious individual and another individual in successive time intervals [*t, t*+*T*),[*t*+*T, t*+2*T*), …, [*t*+(*n*1)*T, t*+*nT*) are independent identically distributed random variables and that *n* is very large. By the Central limit Theorem, the total number of potentially infectious contacts made in [*t, t* + *nT*) has approximately a normal distribution with mean *nx*_0_ and 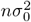, where *x*_0_ and 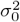 are, respectively, the mean and variance of the underlying distribution in each of the separate time intervals of length *T*. Hence it is reasonable to assume that the total number of potentially infectious contacts has a normal distribution whose mean and variance scale as the total length of the time interval as in our assumptions. Therefore we replace *β*(*t*) in (**??**) by

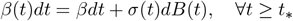

where *B* is a standard brownian motion and *σ* is the diffusion paramter that allows the effect of random control measures.

Imitating the arguments of proof in [], we can show that under the above assumption on *β*, that the underline SDE rewrittes as follows admits a positive solution.

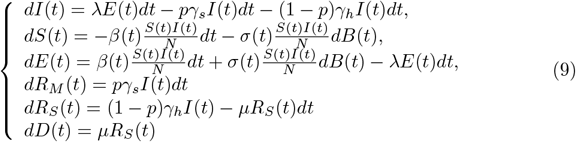

We will performed this section later by adding some relevant pictures according to the value of the diffusion coeficient *σ*.

### 5.2 Conclusion

Finally, let us mention virus mutations that can have a strong influence on the disease progression andtreatment. At the moment, there are no available data on mutations of coronavirus, it may take some time to have confirmation of this aspect. After the universities and schools were closed (March 16) the peak of infection does not seem to be reached, and exponential growth continues. Similar situation is observed in others european nation. This can be an indication that these measures are not sufficient or that they are not respected by local people.

## 6. Appendix

### 6.1 Methods for computing *R*_0_

The basic reproduction number *R*_0_ is defined as the average number of secondary cases generated by a primary case over his/her infectious period when introduced into a large population of susceptibles individuals [**?**]. The constant *R*_0_ thus measures the initial growth rate of the epidemic. A typical first step in analysing a system of differential equations is finding the equilibra. At equilibrium we do not mean that individuals are fixed within particular compartments, but rather the rate of individuals entering and leaving a compartment is excatly balanced. This is equivalent to setting the right hand size of our equations, i.e. the rate of change of individuals into and out of each compartment equal to zero. In epidemiology, models generally have two important equilibria:

- the disease-free equilibrium (DFE) and the endemic equilibrium. The DFE requires that there are no infected individuals in the population, or in the case of SIR model *I*^*\**^ = 0, when the star designates an equilibrium solution. In contrast the endemic equilibrium corresponds to the state in which infected individuals persist indefinitely such that *I*^*\**^ *>* 0.

Finding equilibria is only the first step to understanding long-term behavior in a system. We must also determine which of the behavior are typically realized. This involves determining the stability of the equilibria and will show us whether we will approach the equilibria or more away from it, assuming we have started near by. Now coming back to the method for computing *R*_0_, there is a rich mathematical theory that describe how this quantity can be computed for a long range of SEIR-type models. Let us introduce a commonly used method for finding *R*_0_: the next generation matrix method. of [**?**]. A nice feature of the next generation matrix method is that it only requires use of the DFE which is often easy to compute. In the SEIR model the DFE is given by

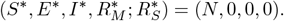

Set

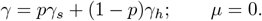

After determining the DFE, we must create a sub-model that only considers the “disease” compartments, a subset of the equations in the modified SEIR model. The disease compartment are those that include individuals that are in any stage of infection which, for the SEIR model, includes both the exposed and infections individuals namely equations:

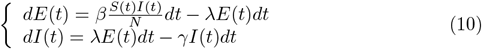

Set

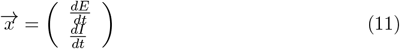

and we can write the disease compartment in the form

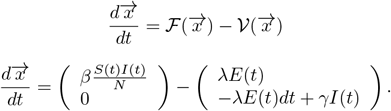

Here 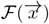 contains any terms that directly lead to new infections entering each compartment *j* (here *j* = 2 because disease compartments are *E* and *I*) and 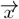 a vector of the *j* disease compartments.

Notice that the second element of ℱ is zero because no new infections enter the compartment *I* rather their transition from to the *E* into *I*.

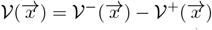

where 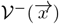 contains all of the outputs and 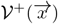 contains all the inputs from each disease class. This includes term such as mortality or transition between class.

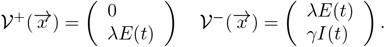

The Jacobian matrix of our equivalent sub-model equations evaluated at the DFE is given by

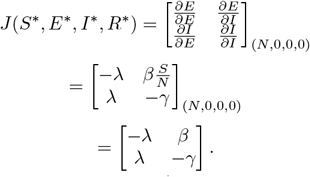

As a result, we can factor out the vector 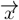 on the right hand size, leaving us with

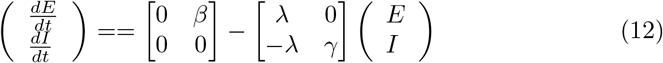

so that

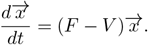

From linear stability analysis we know that the DFE is equivalent to the real part of all eigen values of *F* − *V* being less than zero.

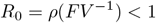

means that the DFE is stable where *ρ* is the spectral radius of *F V* ^−1^.

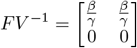

so that

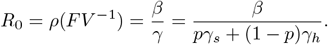

### 6.2 Towards a generalized continuous time Markov SEIR model

The analogous stochastic model (continuous timeMarkov chain) is constructed by considering the following events: exposure *E*, infection *I* and removal (mild or severe)*R*. The transition rates are defined as

| Event | Effect | Transition rate |
| --- | --- | --- |
| Exposed $E$ | $(S, E, I, R) \rightarrow (S - 1, E + 1, I, R)$ | $\beta(t) \frac{SI}{N}$ |
| Infectious $I$ | $(S, E, I, R) \rightarrow (S, E - 1, I + 1, R)$ | $\lambda E$ |
| All Removed | $(S, E, I, R) \rightarrow (S, E, I - 1, R + 1)$ | $(p\gamma_s + (1-p)\gamma_h)I$ |

When an infection occurs, the number of suscptibles decreases by 1 and the number of exposed increases by 1; when the latency period *γ* = (*pγ*_*s*_ + (1−*p*)*γ*_*h*_) ends, the number of number of exposed decreases by 1 and the number of infectives increases by 1; and finally when there is a recovery, the number of infectives decreases by 1, and finally when there is a recovery, the number of infectives decreases by 1 and the number of recovered increases by 1.

increments distributed exponentially:

Now, to understand the following probabilistic modelisation, assumes first that the latency period *λ* and the duration of infection *γ* satisfies an exponential distribution *E ε* (*λ*), *I ε* (*γ*) whe the mean retired rate *γ* = *pγ*_*s*_ + (1 − *p*)*γ*_*h*_. In that case, the above three types of events happen as follows:

- Infection of a susceptible (such an event decreases *S*(*t*) by one, and increases *I*(*t*) by one, so the event happen at rate:

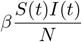
- Exposed (contact) of a susceptible with an infective (such an event increases *E*(*t*) by one, and decreases *S*(*t*) by one, so the event happen at rate:

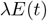
- Removed of an infective with mild infection *R*_*M*_ (such an event decreases *I*(*t*) by one and increases *R*_*M*_ (*t*) by 1 so the event) happen at rate

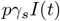
- Retired *R*_*S*_ with severe infection (such an event decreases *I*(*t*) by one and increases *R*_*S*_(*t*) by 1 so the event) happen at rate

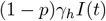

Hence we have the following equations with *P*_*s,e*_, *P*_*e,i*_, *P*_*i,rs*_, *P*_*i,rh*_ *P*_*rh,d*_ standard mutually independent poisson processes:

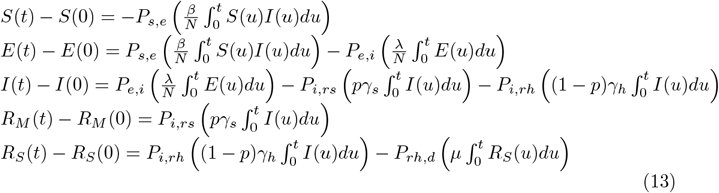

Note that one can clearly forget about the last equation since If we set

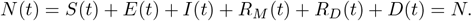

Now define

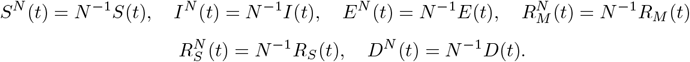

We have

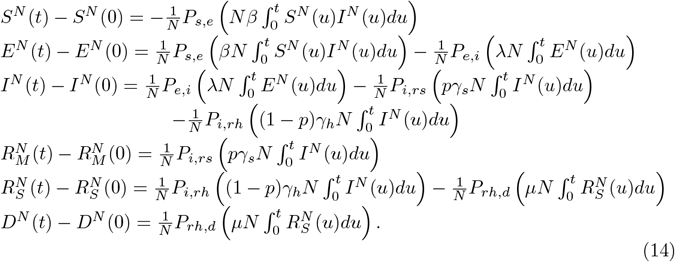

The above model equations assumes that *β, γ*_*s*_, *γ*_*h*_ and *λ* are constant. But as explained above a temporally varying parameter is relevant in order to explained stochastic or deterministic fluctuations in *β* so we consider *β*(*t*) and the model equations can be read as follows.

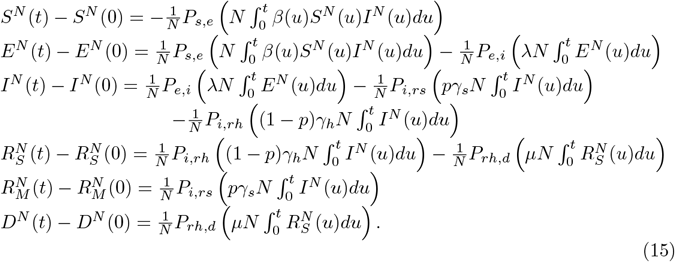

Note that here again the population is closed:

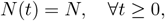

and the above stochastic model can reads as follows:

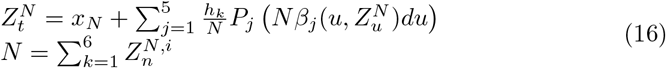

where

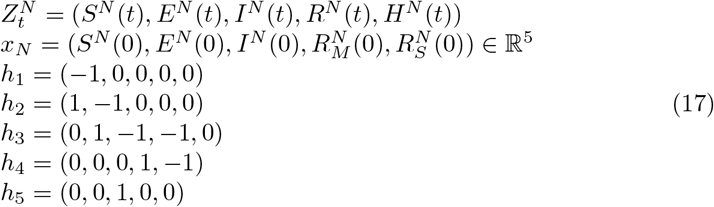

and

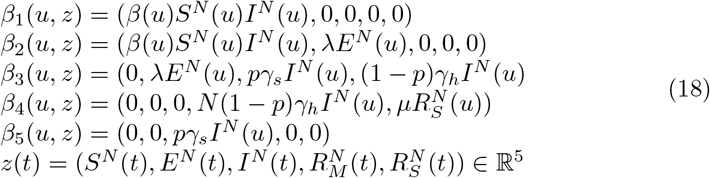

The *P*_*j*_ are standard mutually independent poisson processes defined above.

Note that we forgot about the last equation in *D*(*t*) since it is not relevant in the study of the previous model equations since it can be deduced easily.

In order to estabilished some relevant results let us recall the following Large Numbers for Poisson processes.

**Proposition 6.1** *Let* (*P* (*t*))_*t*≥0_ *be Poisson process with rate λ. Then*

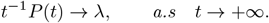

Imitating the arguments in Theorem 2.2.7 in [**?**], we can established the following Theorem.

#### Theorem 6.1

*Law of Large Numbers Assume that*

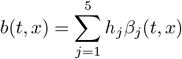

*is a locally Lipschitz function in x, locally uniformly in time t such that*

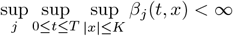

*and the unique solution of the ODE*

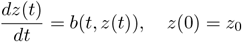

*does not explose in finite time. Let* 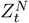 *denote the solution of the above Stochastic Differential Equation. Then* 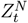 *converges almost surely and locally uniformly in t to*

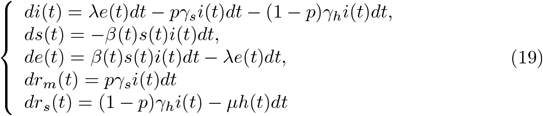

*which is exactly our deterministic model normalized with N with s* = *S/N, i* = *I/N, e* = *E/N r*_*m*_ = *R*_*M*_ */N, r*_*s*_ = *R*_*S*_*/N and z* = (*s, e, i, r*_*m*_, *r*_*h*_).

Now, since the above stochastic model converges to our deterministic diffential system model, it is natural to look at fluctuations of the difference between the stochastic epidemic process and its deterministic limit.

In this way, we introduce the following rescaled difference between 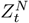 and *z*(*t*):

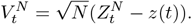

Define the continuous time martingales

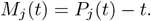

We have

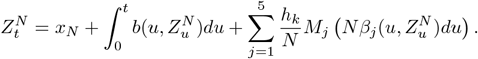

Now, consider the following 5-dimensional process whose *j*-th component is defined as

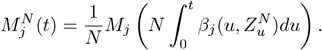

We wish to show that 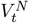 converges in law to a Gaussian process. It is clear that

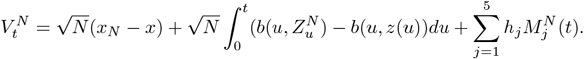

Now let us established the main result of this section. The proof is contained in Theorem 2.3.2 in [**?**].

#### Theorem 6.2

*Central Limit Theorem Under the assumption of the previous theorem, assume furthermore that the function b is locally of class C*^1^ *in x, locally uniformly in t. Then as N→* +*∞ the above rescaled process* 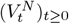 *converges in law to a Gaussian process V defined by the following stochastic differential equation*.

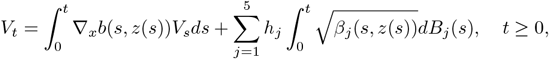

*where* (*B*_1_, *B*_2_, *B*_3_, *B*_4_, *B*_5_) *are mutually independent Brownian motion*.

## Data Availability

https://www.mayotte.ars.sante.fr/coronavirus-covid-19-point-de-situation-et-conduite-tenir

https://www.mayotte.ars.sante.fr/coronavirus-covid-19-point-de-situation-et-conduite-tenir

